# Diabetes Detection from Diabetic Retinopathy-Absent Images Using Deep Learning Methodology

**DOI:** 10.1101/2023.07.27.23287515

**Authors:** Yovel Rom, Rachelle Aviv, Gal Yaakov Cohen, Yehudit Eden Friedman, Zack Dvey-Aharon

**Affiliations:** AEYE Health Inc., Tel Aviv, Israel; The Goldschleger Eye Institute, Sheba Medical Center, Tel Hashomer, Israel; Sackler Faculty of Medicine, Tel Aviv University, Tel Aviv, Israel; Division of Endocrinology, Diabetes and Metabolism, Sheba Medical Center, Ramat Gan, Israel

## Abstract

**Aims:** Diabetes is one of the leading causes of morbidity and mortality in the United States and worldwide. This research aimed to develop an artificial intelligence (AI) machine learning model which can detect the presence of diabetes from fundus imagery of eyes without diabetic eye disease.

**Methods:** Our researchers trained a machine learning algorithm on the EyePACS dataset, consisting of 47,076 images. Patients were also divided into cohorts based on disease duration, each cohort consisting of patients diagnosed within the timeframe in question (e.g., 15 years) and healthy patients.

**Results:** The algorithm achieved 0.83 area under receiver operating curve (AUC) in detecting diabetes per image, and AUC 0.86 on the task of detecting diabetes per patient.

**Conclusion:** Our results suggest that diabetes may be diagnosed non-invasively using fundus imagery alone. This may enable diabetes diagnosis at point of care, as well as other, accessible venues, facilitating the diagnosis of many undiagnosed people with diabetes.

## Introduction

Instances of diabetes, one of the leading causes of morbidity and mortality in the United States, are rapidly increasing and projected to continue climbing. Worldwide, an estimated 536.6 million were living with diabetes in 2021, with an anticipated 46% increase by 2045.^1^ As of 2019, it is estimated that a total 37.3 million (11.3%) Americans have diabetes,^2^ with projections placing the prevalence of diabetes in 2031 at roughly 14%.^3,4^ As a whole, diabetes patients report much higher rates of medical disability than those of the general American population, including disabilities resulting from or comorbid with diabetes.^2^ Among these are vision disabilities, cardiovascular disease, lower-extremity amputation, and chronic kidney disease.^2,5^ While many of these are preventable or may be mitigated with proper care, they are rarely reversible and may develop before treatment is begun.^6^ As such, early diagnosis is critical to the disease management and continuing health of many Americans. Fasting glucose and glycated hemoglobin blood test screening are currently the most established and accepted tools for the diagnosis of diabetes. While both have been shown to be accurate,^7,8^ an estimated 23% of adults with diabetes in the United States and 44.7% of the global adult population with diabetes remain undiagnosed.^2,9^

The eye, and the retina in particular, is a convenient and accessible window into the body in that it is the only place where neural, vascular, and connective tissues may be viewed non-invasively. Much like ocular disorders visibly manifest in the eye, systemic disorders may have manifestations in the fundus, allowing the detection, diagnosis, and monitoring of these diseases. Recent work has shown that even “healthy” fundus images, i.e., images with no humanly diagnosable disorders, can be informative and predictive when presented to a machine learning algorithm. Artificial intelligence (AI) algorithms have shown the ability to interpret subclinical information from retinal anatomy in order to make predictions about systemic indications, such as cardiovascular risk factors,^10^ biomarkers including muscle mass and height,^11^ and chronic kidney disease (CKD) and diabetes.^12^ Additionally, a pivotal FDA study by our group reported on clinical validation for the diagnosis of diabetic retinopathy from a single image per eye, with sensitivity and specificity both significantly above 90%.^13^

One of the key arguments in favor of the use of AI in healthcare is that of scalability.^14–16^ Records may be reviewed, data aggregated, and patients diagnosed in the time it would typically take a doctor to see a patient. Additionally, the portability of many AI systems allows patients access to healthcare in areas or locations that are typically underserved or unvisited by specialists. Although a lack of stable computational infrastructure and resources remains a challenge in particularly low-resource areas and settings, AI systems which utilize smartphones and handheld cameras have been shown to be effective.^16,17^ More specifically, one study showed impressive results with the use of fundus images for the detection of T2DM.^12^

The aforementioned work, which was performed on a Chinese population, utilized both fundus cameras and smartphones in order to accurately diagnose diabetes from fundus images. However, due to the demographically narrow population, and the lack of filtering of patients with diabetic eye disease, there were limitations which this study aims to address. Additionally, in the previous study there was no differentiation between different disease durations, while the patients in the current study were selected for their recent diagnosis and low disease duration. The diagnosis of diabetes is significantly easier and more accurate in patients with a longer disease duration due to vascular changes in the eye, and in this study the accuracy of diagnosis improved in correlation with disease duration.

This study utilized retinal fundus images of patients with and without diabetes, all of whom had no evidence of diabetic eye disease. It aimed to develop an AI system for the early diagnosis of diabetes from the analysis of retinal fundus images.

## Methods

### Dataset

We utilized a dataset compiled and provided by EyePACS (http://www.eyepacs.org), comprised of fundus retinal images. The data consisted of 51,394 images from 7,606 patients who visited the clinics between 2016 and 2021. Of the patients, 39% were male and 61% were female or other; mean age was 56.18 years old (table 1). All images and data were de-identified according to the Health Insurance Portability and Accountability Act “Safe Harbor” before they were transferred to the researchers. Institutional Review Board exemption was obtained from the Sterling Independent Review Board.

**Table 1:** Key dataset characteristics – general.

|  |  |
| --- | --- |
| Number of patients | 7,606 |
| Number of images | 51,394 |
| Age: mean, years (SD) | 56.51 (12.42), n=7605 |
| Gender (% male) | 38.3%, n=7322 |
| HbA1c: mean, % (SD) | 7.82 (2.60), n=4585 |
| Ethnicity | 52.2% Latin American, 13.2% ethnicity not specified, 12.7% Caucasian, 8.8% African Descent, 6.5% Indian subcontinent origin, 3.7% Asian, 1.5% Other, n=7449 |

The dataset contained up to 6 images per patient per visit: one macula centered image, one disk centered image, and one centered image, per eye. Patients’ metadata contained a self-reported measure regarding the presence or absence of diabetes (for demographic data of the two groups see table 2). Images without this metadata were excluded. Indications of diabetic eye disease were provided by professional ophthalmologists as part of the EyePACS dataset metadata and were subsequently excluded as well. Patients were also divided into cohorts based on disease duration, each cohort consisting of patients diagnosed within the timeframe in question (e.g., 15 years) and healthy patients (for demographic data divided by disease duration see appendix A).

**Table 2:** Key dataset characteristics divided by diagnosis.

|  | Patients with diabetes | Healthy patients |
| --- | --- | --- |
| Number of patients | 6,934 | 708 |
| Number of images | 47,076 | 4,318 |
| Age: mean in years (SD) | 56.59 (12.32), n=6934 | 55.85 (13.30), n=707 |
| Gender (% male) | 38.4%, n=6723 | 37.1%, n=634 |
| HbA1c: mean % (SD) | 7.93 (2.52), n=4344 | 5.91 (3.09), n=258 |
| Ethnicity | 51.4% Latin American, 13.2% Caucasian, 12.1% ethnicity not specified, 9.2% African Descent, 7.1% Indian subcontinent origin, 4.0% Asian, 1.4% Other, n=6792 | 60.8% Latin American, 23.2% ethnicity not specified, 7.6% Caucasian, 4.6% African Descent, 2.5% Other, 1.0% Asian, n=693 |

### Algorithm Development

In order to accurately measure performance, given the relatively small size of the dataset, the models were trained using 10-fold validation, i.e., the dataset was divided into tenths and each model was trained on 9/10 parts, and validated on the final tenth. Each model was validated using a different tenth of the data. The model was then validated on the entire dataset.

Additionally, the model was trained on cohorts of patients divided according to disease duration. Each cohort consisted of patients diagnosed within the timeframe in question (e.g., 15 years) and healthy patients. Ten-fold validation was similarly done on these cohorts. Patient images were additionally adjusted for camera type, meaning an equal number of patients with and without diabetes were used across camera types, and age-adjusted for equality between types of patients. Cohort distribution is available in table 1.

## Results

The model’s performance in diagnosing diabetes from fundus images was AUC 0.83 (0.83, 0.84 - 95% CI) per image, and AUC 0.86 (0.84, 0.88 - 95% CI) per patient (see figure 1). The results were unaffected by ethnicity (see table 4) and were affected by camera model (see table 5). For extended versions of these tables see appendices C and D. Model performance also improved as a function of disease duration (correlation 0.92 (p < 0.001), see figure 2). For full model performance statistics divided by disease duration see Appendix B.

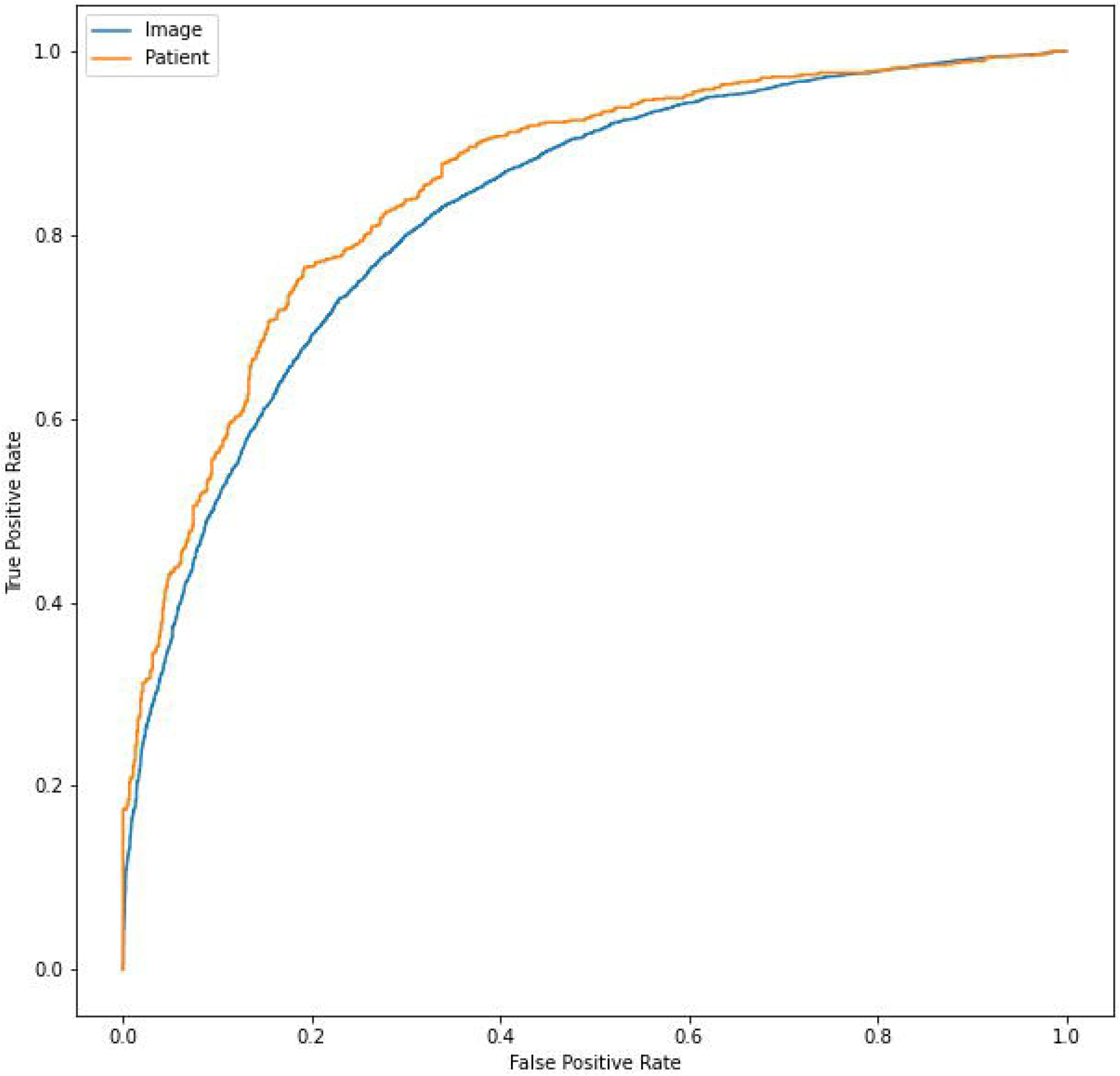

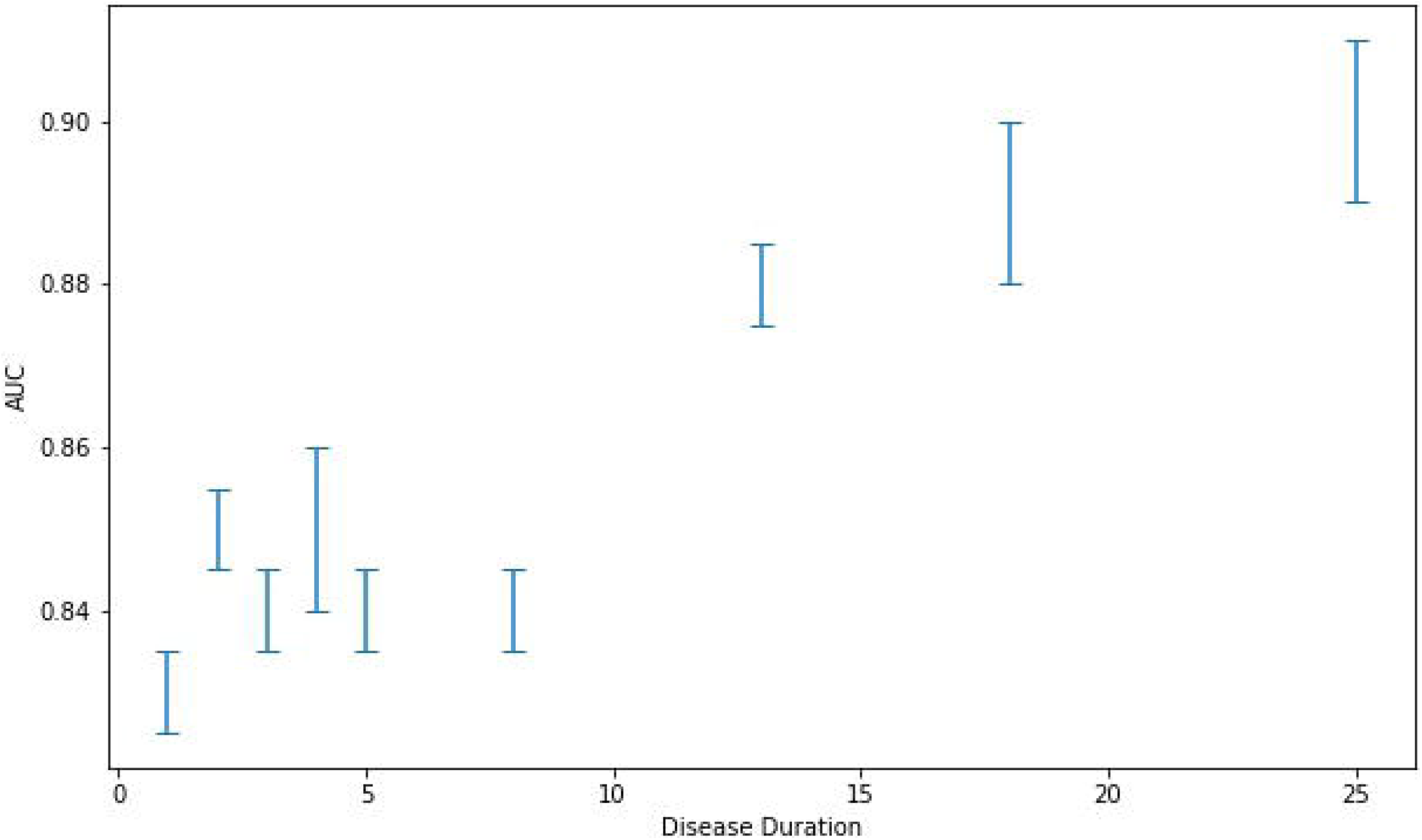

**Table 3:**
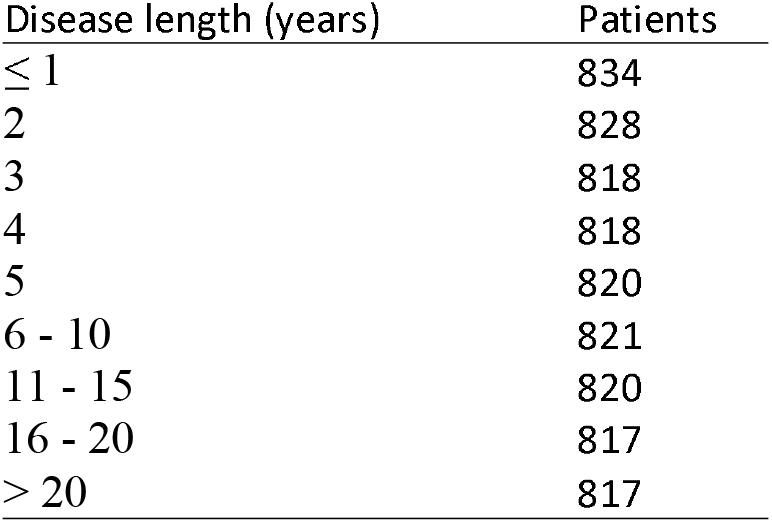
Disease length cohort distribution in years.

**Table 4:** Ethnicity distribution and associated AUC.

| Ethnicity | Patients | AUC | Sensitivity | Specificity |
| --- | --- | --- | --- | --- |
| Ethnicity not specified | 288 | 0.85 (0.80, 0.89) | 0.81 (0.73, 0.87) | 0.70 (0.63, 0.77) |
| Latin American | 901 | 0.84 (0.82, 0.87) | 0.87 (0.84, 0.90) | 0.65 (0.61, 0.70) |
| Caucasian | 163 | 0.87 (0.79, 0.92) | 0.93 (0.87, 0.97) | 0.52 (0.39, 0.64) |
| Other | 37 | 0.70 (0.49, 0.85) | 0.88 (0.62, 1.00) | 0.50 (0.26, 0.70) |
| Asian | 47 | 0.73 | 0.93 (0.80, 0.98) | 0.29 (nan, nan) |
| African Descent | 95 | 0.85 (0.75, 0.91) | 0.90 (0.80, 0.96) | 0.50 (0.32, 0.66) |

**Table 5:** Camera distribution and associated AUC.

| Device | Images | AUC (CI) | Sensitivity | Specificity |
| --- | --- | --- | --- | --- |
| Canon CR2 | 6540 | 0.84 (0.83, 0.85) | 0.82 (0.81, 0.83) | 0.72 (0.70, 0.74) |
| Centervue DRS | 1728 | 0.88 (0.86, 0.89) | 0.88 (0.86, 0.90) | 0.73 (0.70, 0.76) |
| Crystalvue | 498 | 0.51 (0.46, 0.56) | 0.49 (0.42, 0.55) | 0.51 (0.45, 0.57) |
| Topcon NW400 | 592 | 0.58 (0.54, 0.63) | 0.68 (0.63, 0.73) | 0.41 (0.35, 0.47) |

## Discussion

Diabetes is currently one of the leading causes of morbidity and mortality, with many patients remaining undiagnosed. Although diabetes is often diagnosed from fundus imagery after the development of related retinal complications, we hypothesized that a machine learning model could be trained to accurately diagnose patients from images of fundi without diabetic eye disease, doing such before an ophthalmologist can do so. The model performed as expected, further improving as a function of disease duration, seemingly due to the increased impact of the disease on optical microvasculature over time. As previously mentioned, AI-based screening for various conditions offers a low-cost, accessible, and scalable tool with great potential. In the case of diabetes, specifically, the use of AI-based retinal screening may increase testing, due to factors of both comfort and ease. Additionally, due to the same factors, this may also increase early testing which is critical for previously discussed reasons.

Currently, to perform the standard blood work required for screening, a referral from a physician is required. The increased burden of follow-up, or a lack of concern, on the patient’s part may very well be a major cause for under-detection, especially if they are presumably healthy. Many patients remain undiagnosed up to the point of vision impairment and their subsequent ophthalmological exam.^19^

As a whole, novel non-invasive technologies may be of value as alternative screening tools, facilitating early detection and diagnosis of diabetes. There are currently a handful of devices that allow non-invasive detection of diabetes, such as the Scout DS,^20,21^ which uses light to detect advanced glycation end-products (AGEs) and other biomarkers in the skin. To our knowledge, the usage of these technologies in the clinical field is limited.

Novel methods will not necessarily replace traditional ones, but their adoption will enable the development of parallel, physician independent screening processes. Integrating these novel technologies will make screening at accessible sites possible. Possible screening sites could be at workplaces, at pharmacies, shopping malls or even at home, using portable devices. Increasing the screening ratio, and specifically at earlier stages of diseases, may decrease long term microvascular complications,^22^ and may prove to be cost-effective.

There are, however, legal and ethical issues concerning the use of these new technologies. Community screenings, independent of an established healthcare setting, are currently generally not encouraged by the ADA. There are questions of liability on the results of the tests, follow-up testing, and the provision of proper explanations to and treatment of the patients. Lastly, there are economic issues regarding the costs of these tests, coverage by medical insurance companies both for the test itself and further diagnostic tests, and the ability of existing healthcare systems to accommodate the increase in referrals of newly diagnosed patients.

There are numerous ways to mitigate the above-mentioned risks of the “democratization of screening”. In order to properly allow this shift, a comprehensive, interdisciplinary thinking process is required. While further risk-benefit analysis is required, the benefits may very well outweigh the risks, enabling the diagnosis and proper care of millions.

## Limitations and further research

This study’s main limitation, which we hope to address in the future, lies in the research’s retrospective nature, meaning that both prospective research and external validation are lacking. Furthermore, results were affected by camera model, probably due to uneven distribution of camera models in the training set. An additional limitation is that patients’ diabetes status was self-reported, which may have had the effect of lowering the model’s final AUC. Given the extremely high percentages of undiagnosed patients, there is a substantial likelihood that there are people with diabetes among the designated image set of healthy patients. As such, the model’s actual results may be different and more accurate. This research also does not differentiate between type 1 and type 2 diabetes. The likelihood of this differentiation causing significantly different results is low, given the relatively low prevalence of T1DM,^1^ but are still worth exploring. These issues may be possible to address in further research, including prospective research or research utilizing different datasets. It should be noted, however, that research differentiating between T1DM and T2DM would require a much wider pool of patients, including children, in order to include patients with similarly low disease durations between groups.

The diagnosis of pre-diabetes is not included in the scope of the current research, however will be included in future validation of the model in question.

Our work raises an additional clinical question: assuming that machine learning models for the detection of diabetes from retinal images are mainly based on subclinical changes in the vasculature of an end organ, further research would be necessary to define whether medical treatment should differ in patients with positive retinal screening tests or differ based on the model’s degree of certainty in the diagnosis.

## Data Availability

Data may be obtained from a third party and are not publicly available. De-identified data used in this study are not publicly available at present. Parties interested in data access should contact JC for queries related to EyePACS. Applications will need to undergo ethical and legal approvals by the respective institutions.

## Notes

### Competing Interest Statement

YR and RA are employees of AEYE Health. ZD-A is CEO of AEYE Health.

### Funding Statement

This study was funded by AEYE Health, Inc.

### Author Declarations

Institutional Review Board exemption was obtained from the Sterling Independent Review Board.

